# Novel visualization of the spatiotemporal relationship between ictal spiking and LFP supports the involvement of mid-range excitatory circuits during human focal seizures

**DOI:** 10.1101/2022.05.06.22274768

**Authors:** Somin Lee, Sarita S. Deshpande, Edward M. Merricks, Emily Schlafly, Robert Goodman, Guy M. McKhann, Emad N. Eskandar, Joseph R. Madsen, Sydney S. Cash, Michel J.A.M. van Putten, Catherine A. Schevon, Wim van Drongelen

**Affiliations:** Department of Pediatrics, University of Chicago, Chicago, IL 60637; Medical Scientist Training Program, University of Chicago, Chicago, IL 60637; Department of Neurology, Columbia University, New York, NY 10032; Graduate Program in Neuroscience, Boston University, Boston, MA 02215; Department of Neurosurgery, Lenox Hill Hospital, New York, NY 10075; Department of Neurological Surgery, Columbia University, New York, NY 10032; Department of Neurosurgery, Massachusetts General Hospital and Harvard Medical School, Boston, MA 02114; Nayef Al-Rodhan Laboratories for Cellular Neurosurgery and Neurosurgical Technology, Massachusetts General Hospital and Harvard Medical School, Boston, MA 02114; Department of Neurosurgery, Brigham and Women’s Hospital and Harvard Medical School, Boston, MA 02115; Department of Neurology, Massachusetts General Hospital and Harvard Medical School, Boston, MA 02114; Clinical Neurophysiology Group, MIRA Institute for Biomedical Engineering and Technical Medicine, University of Twente, Enschede 7500AE, The Netherlands

**Author notes:** Corresponding Author: Wim van Drongelen. Designed research: S.L., S.S.D., M.J.A.M.v.P., C.A.S., E.M.M., W.v.D. Performed research: S.L., S.S.D., E.M.M., E.S., E.N.E., J.R.M, S.S.C., R.M., G.M.M., M.J.A.M.v.P. W.v.D. Analyzed data: S.L., S.S.D., W.v.D. Wrote and edited manuscript: S.L., S.S.D., E.M.E., J.R.M., S.S.C., M.J.A.M.v.P., C.A.S., W.v.D. S.L and S.S.D. contributed equally to this work. The authors declare no competing interests.

**Keywords:** Focal seizure activity, Mid-range connectivity, Spatiotemporal spike-centered average, Ictal local field potential, Spike-triggered average

## Abstract

The relationship between action potentials and the associated local field potential (LFP) in neural recordings is typically studied only in the temporal domain using the spike-triggered average (STA). In this study, we present a novel approach, termed the spatiotemporal spike-centered average (st-SCA), that allows for visualization of the spike-LFP relationship in both the temporal and spatial domains. In this method, a 3D spatiotemporal topography of spike-associated LFP is calculated from a 2D spatial average of the LFP centered around the time and location of individual spikes. We applied this method to 25 microelectrode array (MEA) recordings obtained from seven patients with pharmacoresistant focal epilepsy during ictal and interictal states. Five patients in this dataset had MEA implants in recruited cortex, and two had implants in unrecruited cortex. Of the five patients with arrays implanted in recruited territory, three showed STAs that resembled sine cardinal (sinc) functions, and two showed non-sinc functions. Using the st-SCA, we found that the patients who showed a sinc-function pattern in the temporal domain showed a donut-shaped ring of LFP activity in the spatial domain. This observation was corroborated by a theoretical model describing an ictal spike as measured by a macroelectrode. The model also revealed a special symmetry wherein temporal component of the st-SCA predicts the spatial component when they both approximate sinc-functions. Supporting this theoretical derivation, a radial cut of the donut-shaped st-SCA showed a spatial pattern consistent with a sinc-function. This spatial sinc-function had peaks separated by ∼2.5mm—a measurement that supports the role of mid-range excitatory connections during ictal activity. In sum these findings suggest that patients whose seizures engage mid-range connections may be identifiable by the spatiotemporal features of ictal spike-associated LFP activity.

## Introduction

Spatiotemporal patterns of brain electrical activity reflect neural mechanisms underpinning different brain pathologies. Consequently, temporal and spatial patterns observed in electrographic recordings are frequently employed to guide diagnostic and therapeutic approaches in the treatment of epilepsy. During surgical evaluation of patients with epilepsy, a variety of electrodes are used to record brain electrical activity across different scales. For example, large-scale global activity can be recorded by macroelectrodes at the scalp or cortex, and meso- and microscale activity can be recorded by intracranial arrays or bundles of microelectrodes (Eissa et al., 2017; Eissa et al., 2016; Schevon et al., 2012). Despite the heavy reliance on electrophysiology in clinical practice, the relationship between neural activity across scales and the mechanistic implications of the observed spatiotemporal patterns remain poorly characterized.

One important question in understanding cortical seizure dynamics is how the activity of individual neurons relates to local and global network activity in ictal and interictal states. The interactions of neural networks during human focal seizures across micro-, meso- and macroscopic scales have been characterized by other recent studies (Eissa et al., 2017). Specifically, one study showed that the spike-triggered average (STA) of the ongoing low frequency component of the local field potential (LFP) could be approximated by a sine cardinal (sinc) function (Eissa et al., 2018). Furthermore, filtering a train of ictal action potentials with a rectangular (brick wall) filter generated an output that correlated well with the observed seizure, consistent with the fact that the Fourier transform of a rectangular function is the sinc-function (Van Drongelen, 2018). While the ictal STA was determined in the temporal and frequency domains, the spatial component of the relationship between action potentials and low frequency LFP was not characterized.

Similarly, most previous studies that describe the relationship between single spiking activity and the surrounding LFP have focused primarily on temporal descriptions using the STA (Bazelot et al., 2010; Glickfeld et al., 2009). The few studies that have investigated the spatial component of this relationship do so by incorporating spatial information into the STA through the addition of spatial filters (Telenczuk et al., 2017) or use a covariance-based approach (Rust et al., 2004). None so far have directly visualized the full spatial topography of LFP associated with spiking activity.

In this study, we present a novel approach, termed the spatiotemporal spike-centered average (st-SCA), in which a mesoscale spatial topography of spike-associated LFP can be visualized by calculating a spatial average of the LFP centered around the location of individual spikes. Calculation of this topography results in a powerful tool that allows for the visualization of both the spatial and temporal components of the spike-LFP relationship. We apply this method to microelectrode array (MEA) recordings of human focal seizures to reveal unique spatiotemporal patterns that support the role of mid-range excitatory connections during ictal activity. We then combine these MEA observations with a mathematical model describing an ictal spike as measured by a macroelectrode to show that in special cases, the temporal and spatial features of the spike-associated LFP can predict one another. In the discussion, we explore the biological mechanisms and clinical implications of the newly observed spatiotemporal properties in the context of pharmacoresistant focal epilepsy.

## Results

To expand upon the traditional temporal characterization of the complex relationship between neuronal firing and the overlying global LFP (Movie S1), we introduce a novel calculation termed the spatiotemporal spike-centered average (st-SCA). The st-SCA builds upon the more typically utilized STA by accounting for both spike timing and location. The st-SCA is determined with respect to the action potential’s timing (*t*, temporal component) and location in the cortical plane (*x, y*; spatial component). To do this, we characterize the multi-unit action potential train (with *i* = 1, …, *N* action potentials occurring at *x*_*i*,_ *y*_*j*,_ *t*_*j*_) as a series of unit impulses:

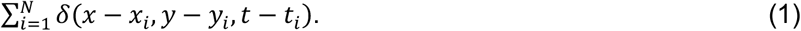

This produces the expression for the normalized spatiotemporal cross-correlation *C*(*ξ, ψ, τ*) between the LFP and action potential:

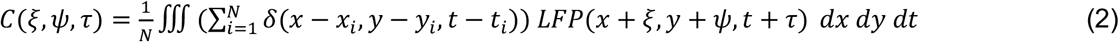

To evaluate this expression, we interchange the integration and summation operations and integrate over the spatiotemporal domain. The resulting expression is defined as the st-SCA:

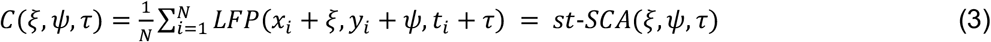

Note that if we set the range of (*ξ, ψ*) equal to the area covered by a fixed spatial range, we obtain the well-known temporal STA (Fig. 1A). In contrast, if we set *τ* to a fixed temporal range, we obtain purely the spatial component of the st-SCA for that epoch. In the following, we describe the computational steps to determine the st-SCA in MEA recordings.

**Figure 1.**
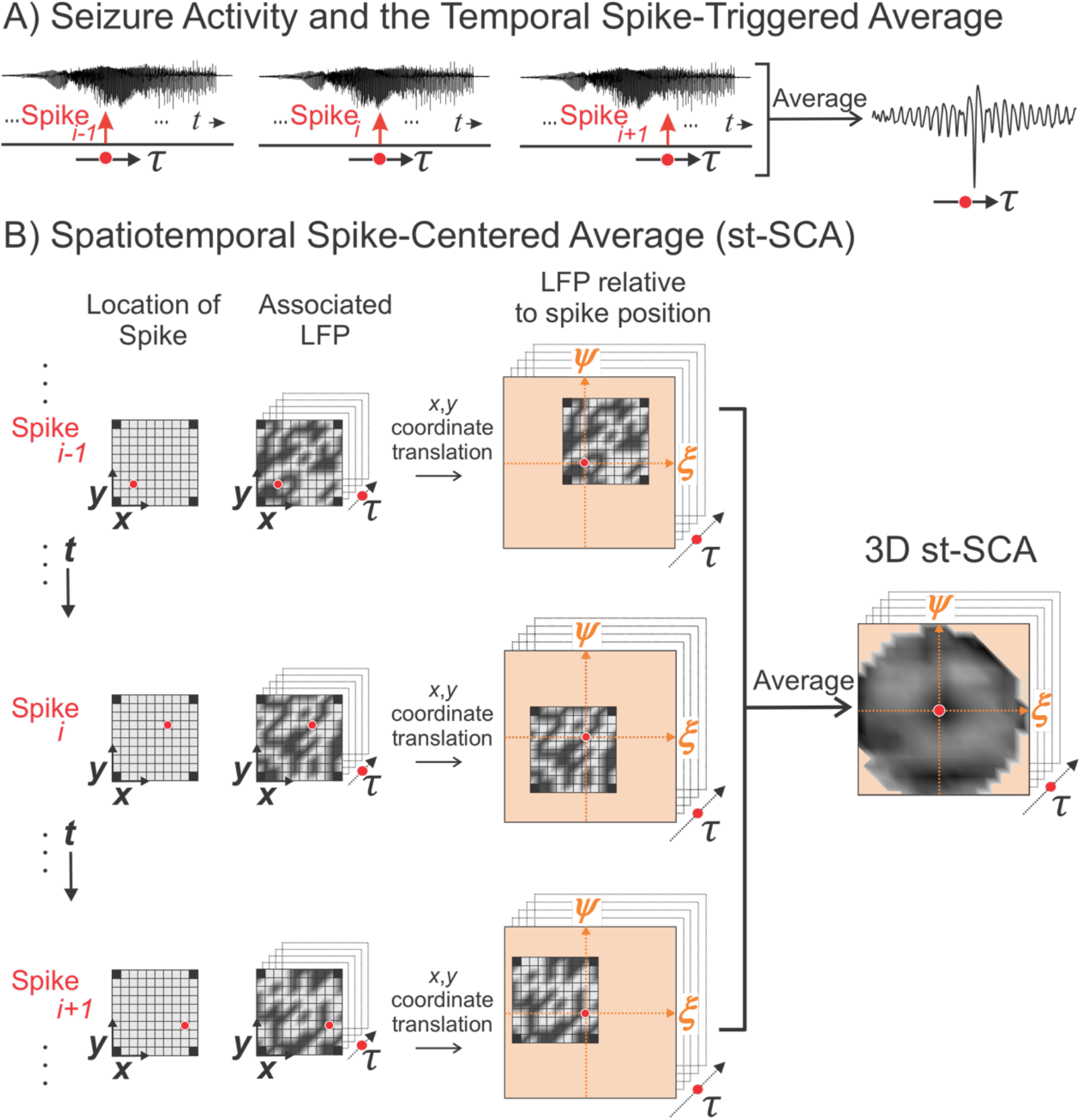
The method to compute the mesoscale spatiotemporal spike-centered average (st-SCA) between spiking activity and the low frequency component of the local field potential (LFP) during a human focal seizure involves centering and averaging each spike’s associated LFP in both time and space. A) During seizure activity, the LFPs within the area of the electrode array (the summed LFP of the microelectrode array is depicted in the upper trace) are associated with a multi-unit action potential train. The LFP’s relationship to the spike is considered over time *τ* relative to the spike events. B) For each spike (left column) across the MEA, its associated spatiotemporal LFP (middle column) is determined. The red circle in the middle column indicates the spike position on the MEA. Next, the (*x, y*)axes of the LFP are translated into the (*ξ, ψ*) axes, such that the associated spike position is at the origin (right column). Finally, the results in the right column are averaged to create a matrix that contains the st-SCA. Note that the corners of the average are undefined because the MEA does not have electrodes in the corner positions.

### Calculation of the st-SCA in MEA recordings

To apply the st-SCA to MEA recordings, we must account for the irregular timing and location of spiking activity across the array. A simplified analogy of this approach is to visualize spiking activity as stones being tossed into water. Consider throwing a single stone into water and analyzing the consequent effects by observing the resulting water ripples. We can simulate multiple sources by dropping identical stones from the same height but at different times and locations across the horizontal plane of the water surface, resulting in a complex landscape. To determine the contribution of a single stone to this landscape, we can take a field of view centered around individual stones. According to Eq. 3, averaging across all stones gives us the spatial pattern of activity associated with each stone. If we also include the time interval around each dropped stone, we obtain the stone’s characteristic spatiotemporal perturbation.

To apply this to the analysis of MEA recordings, spikes are detected for each channel in the MEA (Fig. 1B, left column), and the low frequency LFP associated with each spike is determined (Fig. 1B, middle column). This LFP is then spatially translated such that the associated spike position (*x, y*) is at the origin of a new axes (*ξ, ψ*) (Fig 1B, right column). This spike detection and LFP translation process is then applied to all channels. Averaging the results across all channels results in a field of view of the spike-associated LFP that is (1) centered around individual spikes and (2) approximately four times larger than the area of the MEA (Fig. 1B, right panel). This field is then calculated for time points *τ* to result in the st-SCA.

### Application of st-SCA to clinical recordings allows visualization of spatiotemporal dynamics of human focal seizures

We applied the st-SCA method to analyze microelectrode recordings of focal seizures in patients undergoing epilepsy surgery evaluation. These recordings were obtained from 96-channel, 4×4mm MEA during and around ictal activity (Schevon et al., 2012; Truccolo et al., 2011). Filtering was used to extract local multi-unit neural firing activity and the associated low frequency component of the LFP of the surrounding network (Methods). A total of 19 seizures from seven patients was used for the study (Table S1). Both ictal and interictal recordings were evaluated, where interictal was defined as being at least two hours away from any known ictal activity. Of the seven patients analyzed for this study, five patients had arrays implanted in the recruited seizure territory (Patients 1-5), and two patients had arrays implanted in unrecruited seizure territory but within the clinically-determined seizure onset zone (Patients 6-7) (Schevon et al., 2012). As previously described, recruited seizure territory is defined as local tissue invasion by the seizure wavefront. Unrecruited territory is located outside of the recruited territory but can show rhythmic EEG activity due to local synaptic activity (Merricks et al., 2021; Schevon et al., 2012).

The representative STAs calculated across ictal and interictal states for two recruited territory recordings and one unrecruited territory recording are depicted in Fig. 2. The black lines represent the STAs, the red lines represent the associated noise estimates, and the vertical dotted lines indicate *t* = 0, i.e., the timing of the spike trigger. For both recruited and unrecruited territory recordings, the amplitudes of the ictal STAs (Fig. 2C, E, G) were larger than the corresponding the interictal STAs (Fig. 2D, F, H).

**Figure 2.**
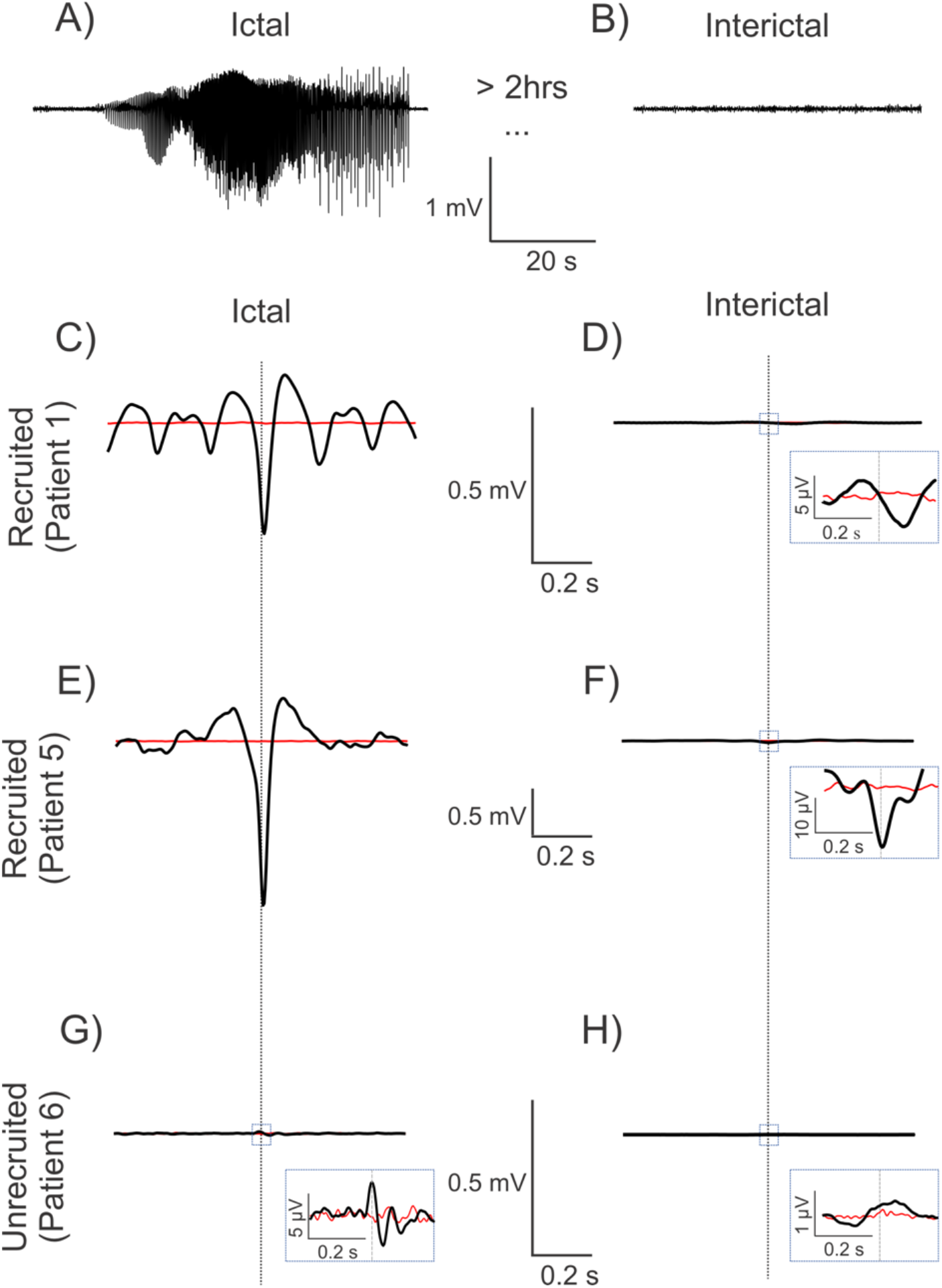
Spike-triggered averages (STAs) in recruited and unrecruited cortical territories during ictal and interictal phases show different patterns. The black traces are the signals, and the red traces represent the associated noise estimates. Vertical stippled lines represent the zero of the time-axis. A—B) Example signal trace of average ictal and interictal LFP activity across MEA channels. C—F) The STA in the recruited territories show an evolution towards a characteristic negative peak, with or without surrounding oscillations, during the ictal phase. The ictal phase amplitudes are also much higher than those of the interictal phase. G—H) The STA in the unrecruited territory show much smaller, non-zero ictal signals than the ones in the recruited territories. In contrast to the recruited territories, the small but dominant ictal peak polarity is positive in this case. The insets in panels D, F, G, H show that unrecruited ictal and across all interictal phases show smaller amplitude, albeit non-zero STAs.

The amplitude for the unrecruited ictal STA (Fig. 2G), however, was much smaller than the recruited ictal STAs (Fig. 2C, E).

Patients with the MEA located in recruited seizure territory showed STAs with different morphologies (Fig. S2A-E), but all had a dominant negative peak around *t* = 0. Consistent with previous findings, we found that the STA for Patients 1-3 resembled a sinc-function with a peak embedded in a weak oscillatory component (Fig. 2C, 3A, S2A-C) (Eissa et al., 2018). In contrast, the STA for Patients 4 and 5 did not resemble a sinc-function as Patient 4 showed a dominant peak embedded in a strong oscillation (Fig. S4D) while Patient 5 showed no oscillatory component (Fig. 2E, Fig. 3B, S4E). The STAs for Patients 6 and 7 with the MEA in unrecruited territory were weak with a smaller amplitude deflection around to *t* = 0 (Fig. 2G, Fig. S4F, G).

**Figure 3.**
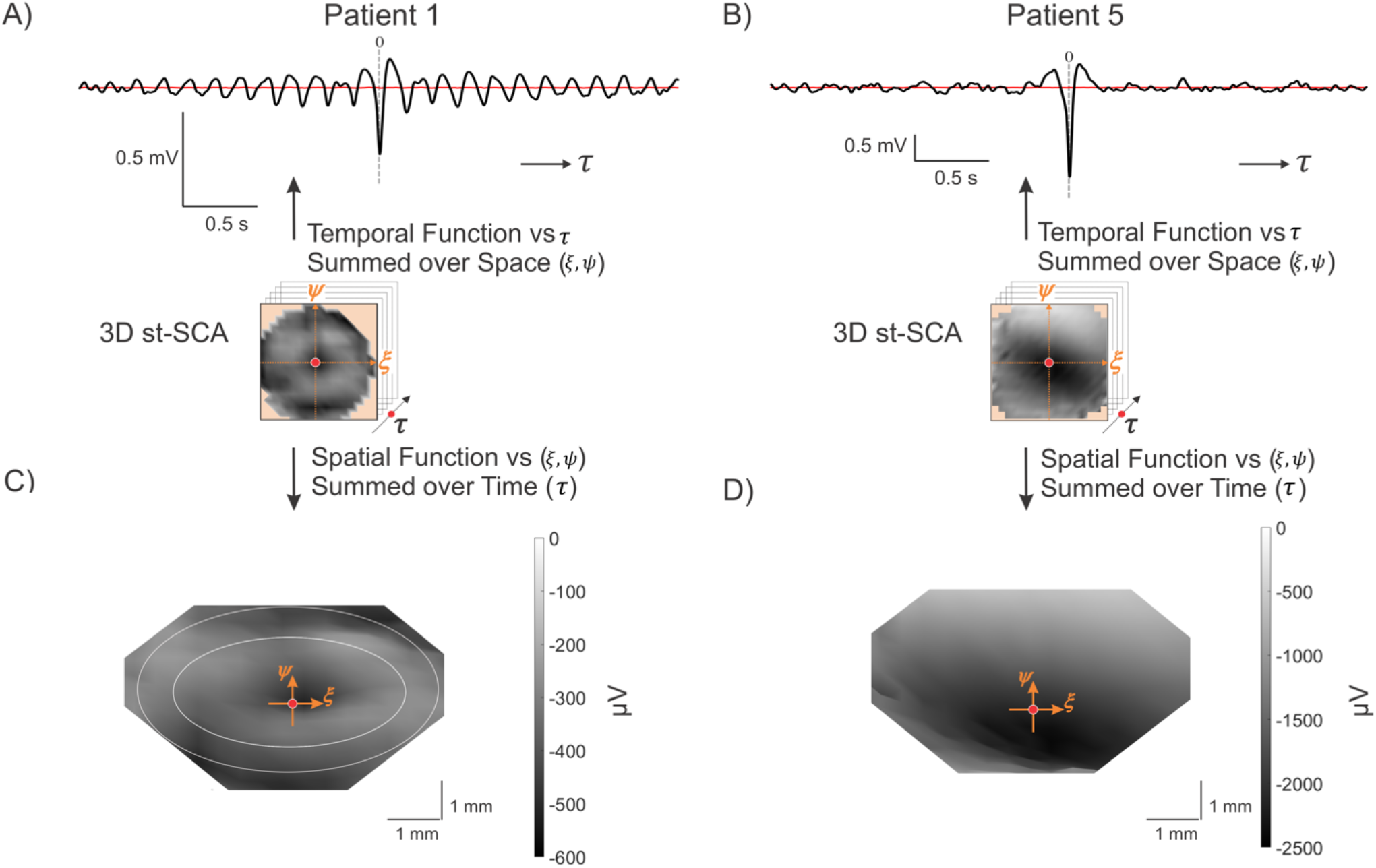
Properties of the ictal spatiotemporal spike-centered average (st-SCA) function during a focal seizure show different patterns for two representative patients. A—B) The temporal average is calculated by averaging the st-SCA over all spatial contributions (±3.6mm). C—D) A 3D view (azimuth = 0°, elevation = 70°) of the 3D st-SCA summed over time *τ* =±35ms. The center (*ξ, ψ* = 0,0) is indicated by the red dot. The two concentric circles are drawn to indicate that the center is surrounded by two rings. Note the apparent radial symmetry of the st-SCA pattern in (C). Grayscale is in μV units.

We then evaluated the relationship between spiking activity and the LFP in the spatiotemporal domain by computing the st-SCA over the entire MEA (*ξ, ψ*) and times *τ* = ±1ms (Fig. 3C, D). This 2ms interval averaged across to yield a 2D spatial topography. In the ictal phase for Patients 1-3, we observed a centrally located trough surrounded by a pair of rings with apparent radial symmetry (Fig. 3C, Fig. S2A). The distance between the center and the region indicated by the inner circle was ∼1.5mm, and the distance between center and the region indicated by the outer circle was ∼2.5mm (Fig. 3C). In contrast, the ictal phase for Patients 4 and 5 showed a deep well of stronger negative activity (e.g., Fig. 3D). The st-SCAs during the interictal phase as well as the results obtained in unrecruited territories showed different patterns with relatively smaller amplitude signals (Fig. S2B, D, E, F). For a list of representative temporal and spatial STAs for each patient, see Fig. S2.

In sum, three of five patients with recordings from recruited territories (Patients 1-3) showed st-SCAs resembling sinc-functions in the temporal domain and st-SCAs resembling donut-shaped rings of activity in the spatial domain. In Patients 4 and 5, the STAs were characterized by a non-sinc morphology, and the st-SCAs showed deep and diffuse wells of negative activity (Fig. 3D, S4D, E).

Note that these observations were not attributable to widespread correlations among MEA electrodes. To demonstrate that the observed st-SCA patterns are representative of the spike-LFP relationship and not the global, macroscale correlations amongst network LFPs, we showed that STAs in unrecruited territory show a large and significant oscillatory component only when triggered by spikes from recruited territories (Fig. S4B), and not when triggered by spikes from unrecruited territories (Fig. S4C). This result is a replication of previous studies (Eissa et al., 2017). Furthermore, randomizing the spike times detected across the MEA resulted in complete destruction of the observed st-SCA patterns, emphasizing the importance of spike timing as the driver for these spatiotemporal patterns (Fig. S5).

Finally, calculation of the st-SCA after applying a spatial filter to decorrelate LFP signals across MEA channels did not qualitatively alter the st-SCA patterns (Methods, Fig. S6).

### Theoretical model reveals a unique symmetry that allows for prediction of temporal and spatial components of the st-SCA

The st-SCA calculations presented above were obtained indirectly by an averaging procedure in order to reduce the noise not associated with the spike trigger in the spike-LFP relationship. In this section, we simulate an ideal noise-free, controlled environment where spatial and temporal relationships can be measured directly (Fig. 4).

**Figure 4.**
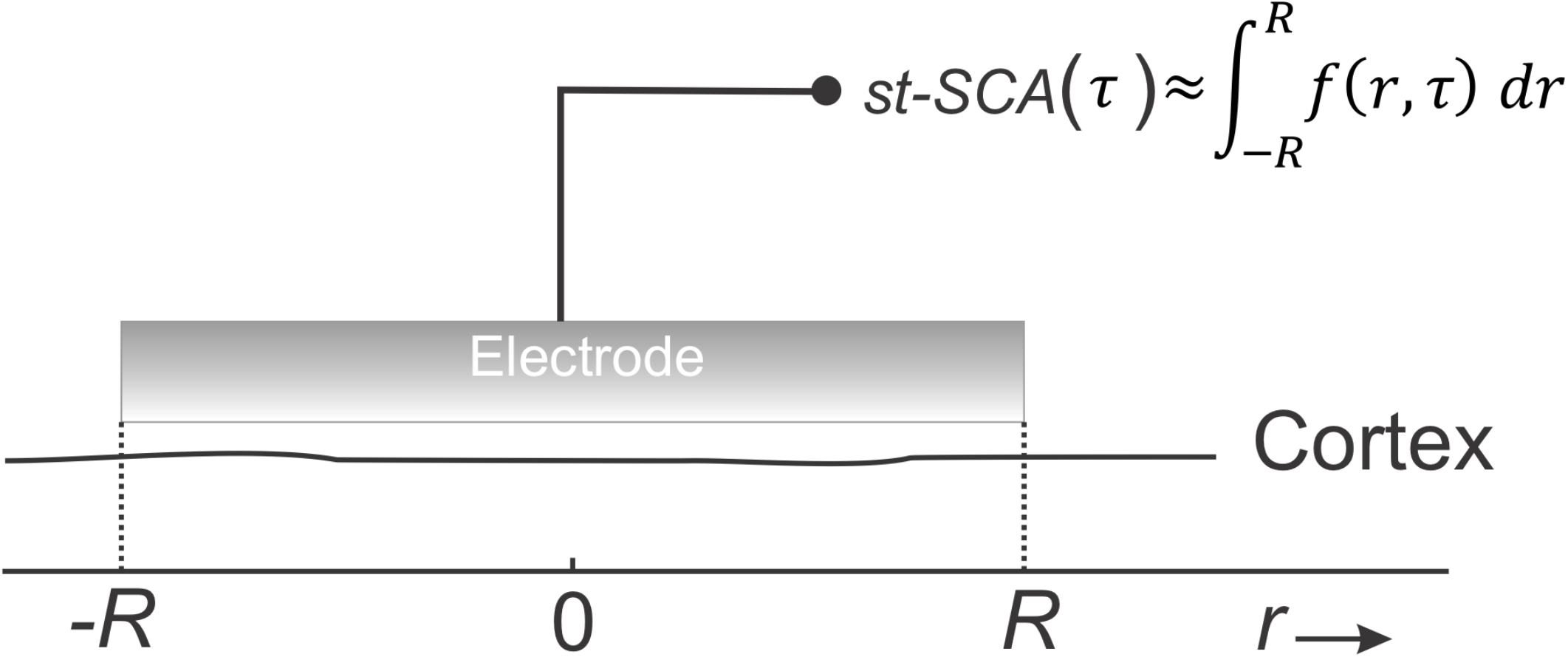
A mathematical model as represented by the recording of macroelectrode measures the underlying network’s st-SCA. The electrode covers an area of one-dimensional cortex where we record the effect associated with a single centrally located ictal action potential, *δ*(r, τ) = δ(0,0), i.e., the macroelectrode measures the underlying network’s temporal component of the st-SCA, *st-SCA*_4_(*τ*). This measurement can be approximated by an unknown action potential’s spatiotemporal cortical activation function, *f*(*r, τ*), integrated over the spatial range [−*R, R*] covered by the electrode.

Here, the sum of all measurements across the MEA can be represented by a single macroelectrode, which records cortical activity when a spike occurs at time zero and at the center of the electrode. This is a measurement of the noise-free spatiotemporal local field function *f*(*r, τ*) (space (*r*) and time (*τ*)) associated with the central spike represented by a delta function, *δ*(*r, τ*). Because the potential of cortical generators attenuates sharply with distance, we consider contributions from activity in areas not directly under the macroscopic electrode to be negligible. Under this assumption, the electrode’s signal can be approximated by summing the contributions over the neocortical area under the electrode, and we find the following expression for *STA*(*τ*):

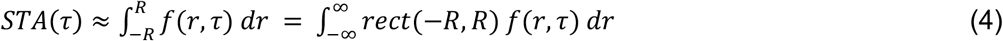

with *rect*(−*R, R*) representing a rectangular window bounded by [−*R, R*]. Similarly, if we compute the spatial component of the SCA, *st-SCA*_*s*_(*r*), by integration over a fixed time epoch, [−*T, T*], around the seizure onset, we get:

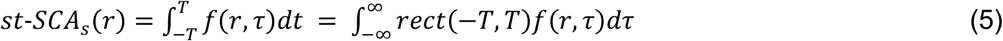

Because the action potential can be represented by a unit impulse, *STA*(*τ*) and *st-SCA*_*s*_(*r*) are equivalent to a cortical unit impulse response (UIR). The UIR can be used to link the spike train to the associated LFP (Supplementary Information). In most cases the function *f*(*r, τ*) cannot be simply derived from measuring the *STA*(*τ*) and *st-SCA*_*s*_(*r*), but one could apply an *ad hoc* solution so that Eq. 4 and 5 produce the correct characterization:

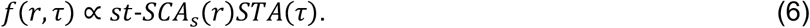

In special cases, however, *f*(*r, τ*) may be mathematically derived. Note that we determined earlier that in three of five patients with recordings in the recruited territory, the *STA*(*τ*) resembles a sinc-function:

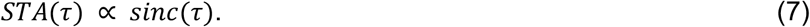

This represents a unique scenario in which the *f*(*r, τ*) and *st-SCA*_*s*_(*r*) may be predicted from the characterization of *STA*(*τ*) alone. The sinc-function is the UIR of an ideal filter, and a previous study has used this property to show that an ictal spike train passed through an ideal filter recreates the seizure’s LFP (Eissa et al., 2018). Since the sinc-function is defined as the Fourier transform of a rectangular function, the relationship between time and space conveniently parallels a time-frequency Fourier-transform-pair. Accordingly, we find:

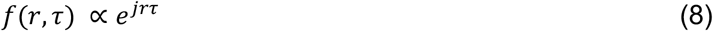

Substitution of this finding into Eq. 5 enables us to find the *st-SCA*_*s*_(*r*), which represents the spatial postsynaptic effects of an action potential effective over a fixed time epoch around the seizure onset ([−*T, T*]):

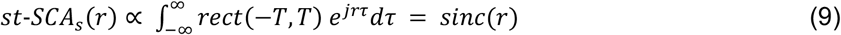

Thus, we find that in the special case where both *STA*(*τ*) and *st-SCA*_*s*_(*r*) are described by sinc-functions, the temporal features of the spike-associated LFP can predict the spatial features, and vice versa.

### Quantification of the peak-to-peak distance of spatial patterns supports the involvement of mid-range excitatory connections

Since the model showed that a sinc-function in the temporal domain predicts a sinc-function in the spatial domain, we aimed to more quantitatively describe the donut-shaped activity observed in Fig. 3C. Taking advantage of the radial symmetry observed in the st-SCA, we converted the Cartesian coordinates (*ξ, ψ*) into polar coordinates (*r, θ*) and focused on the spatial relationship with respect to *r* (Fig. 5A). This enabled us to depict the st-SCA in two dimensions, (*r, τ*) (Fig. 5B). A detail of that relationship is depicted in Fig. 5C, and the summed values across this two-dimensional detail are plotted along its margins. These summed values are the two components as a function of space and time (*r* and *τ*). Note that the bottom graph in Fig. 5C represents the central trough (*τ* = ± 35ms) of the function shown in Fig. 3A. As anticipated by the outcome in Eq. 9 we observed a spatial component (Fig. 5C) that shows a central trough with smaller amplitude side lobes---a pattern consistent with the shape of a sinc-function. Note that the resolution and range of the spatial component (*r* = ±3.6mm) is limited by the size of the MEA (Methods, Fig. 1B). Consistent with the donut-shaped rings observed in Fig. 3C, the peaks of the function shown in the side panel of Fig. 5C were separated by ∼2.5mm (blue arrows, Fig. 5C).

**Figure 5.**
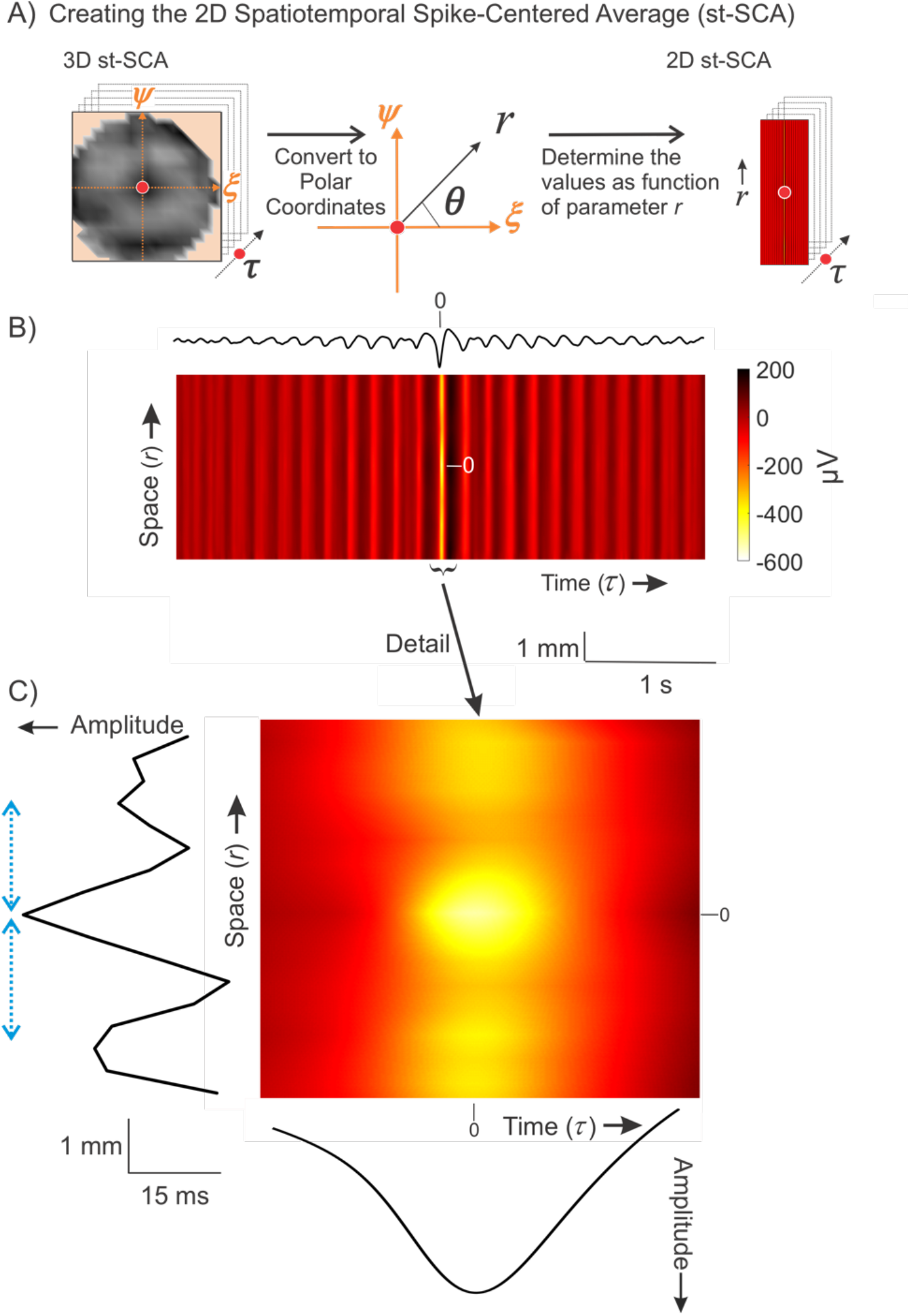
The method to compute the 2D spatiotemporal spike-centered average, *st-SCA*(*r, τ*) allows for a detailed visualization of spatial and temporal components for Patient 1 A) The Cartesian coordinates (ξ, ψ) from the 3D st-SCA are converted into polar coordinates (*r, θ*), resulting in a 2D st-SCA. B) A color representation of *st-SCA*(*r, τ*). The top trace, the temporal component of the *st-SCA*_*t*_(*τ*), is obtained by the sum of *st-SCA*(*r, τ*) over *r* (same as the signal in Fig. 3A). Amplitude and color scale are in μV. C) Detail of the central part of Panel B. The left margin shows the resulting wave from summation over time, generating the spatial component of st-SCA. The green arrows on the left indicate the distance (∼2.5mm) between the peaks seen in this function. The bottom margin depicts the resulting wave from summation over space, generating the temporal component of the st-SCA.

## Discussion

The neural dynamics of human focal seizures show a complex relationship between action potential activity and the LFP (Movie S1). Because characterizing this relationship is important for understanding seizure generation and propagation, this study aimed to determine the spatiotemporal patterns observed across seizure states and cortical locations. Analyses of clinical recordings (Fig. 2, 3, 5) showed that the spike-LFP relationship for some focal seizures can be approximated by a sinc-function in both the spatial and temporal domains. Our theoretical model (Eq. 4-9; Fig. 4) further showed that a sinc-function in the temporal domain can predict a sinc-function in the spatial domain. Here, we discuss the potential biological implications of our findings.

Under physiological conditions, synaptic activity is a major contributor to the extracellular potential field (Nunez et al., 2006). Other contributors may include intrinsic membrane currents, gap junctions, neuron-glia interactions, and ephaptic effects (Buzsaki et al., 2012; Herreras, 2016). While the relative contributions of these different mechanisms during pathological states such as seizures have not been fully elucidated, a non-zero cross-correlation between action potentials and LFPs is expected because synaptic currents are a major component in the compound activities observed in ictal states.

In our discussion of the mechanistic implications of the observed st-SCAs, we assign a net excitation to negative deflections and net inhibition to positive deflections, as previously described (Eissa et al., 2017). Accordingly, our st-SCA analyses (Fig. 2, 3) show that in the recruited ictal territory, the spike-LFP correlation at small lags are dominated by net excitation during seizures in all patients. The activity level in the excitatory center, representing the activity at the ictal wave, is excessively high, possibly due to saturation of the local inhibitory population (Tryba et al., 2019). In Patients 1-3 we also observe a ring of reduced excitation at a distance ∼1.5mm around the excitatory center (Fig. 3C, Fig. S2A). In turn, the ring of reduced excitation is surrounded by a second ring at an additional distance of ∼1mm where excitation increases again. For these patients, this donut-shaped st-SCA is specific to the recruited seizure territory in the ictal phase (Fig. S2). This observation suggests that the ictal wave in the recruited territory, represented by the excitatory center (*ξ, ψ* = 0,0), creates an escape of hyperexcitation via a jump that engages mid-range connectivity in the millimeter range (Fig. 3, 5). Decorrelation of the LFP prior to the st-SCA calculations yielded similar spatiotemporal patterns (Fig. S5), further corroborating the importance of local mm-range excitatory connections in focal seizures.

A question that remains is the biological basis for this connectivity. Histological studies have shown that there are indeed excitatory mid-range connections at the millimeter scale mediated by axon collaterals within the gray matter in the neocortex in addition to short-range excitatory and inhibitory connections at a scale of hundreds of μm (Fig. 6A) (Nieuwenhuys, 1994; Oberlaender et al., 2011; Pichon et al., 2012; Zhang & Deschênes, 1997). Additionally, previous studies of ictal wave dynamics provide direct evidence that mm-range connections are invoked during seizure activity (Schevon et al., 2012). An example of this jump in action potential activity is depicted in the spatial plot in Fig. 6A (a snapshot of Movie S1), in which there are multiple areas of simultaneously increased neural activity across the MEA, separated by mm-range gaps. This is consistent with the distance between the excitatory center and outer ring we observe in the donut-shaped spatial cross-correlation depicted in Fig. 3C. This pathological escape of uncontrolled excitation across cortex could be considered a candidate mechanism in seizure recruitment and propagation (see SI: Mechanisms for Focal Seizures).

**Figure 6.**
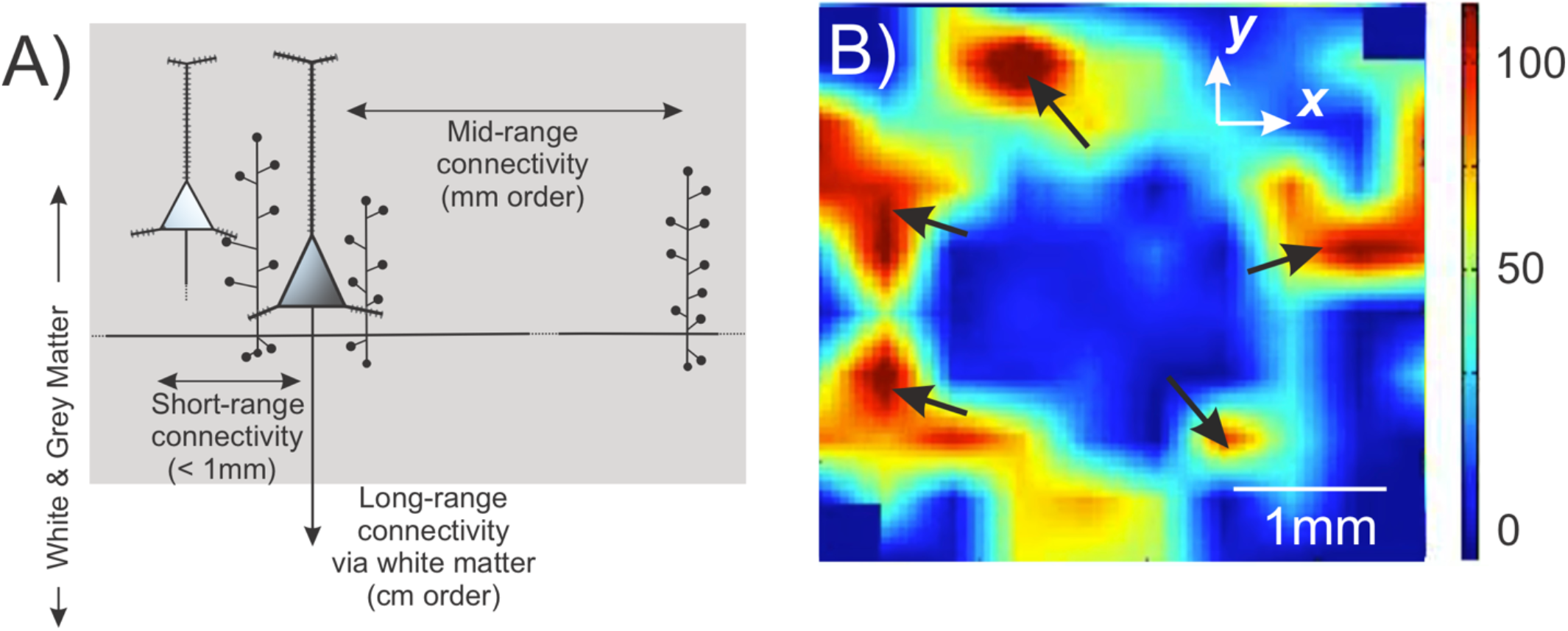
The propagation of the ictal wavefront corroborates the involvement of mid-range excitatory neocortical connectivity by axon collaterals. A) Diagram of gray matter excitatory connections of a neocortical pyramidal cell showing the short-range connections (order of 100s of μm) and mid-range connections (order of mm) via the pyramidal cell axon collaterals (based on Fig. 5 in Nieuwenhuys, 1994). B) Snapshot of Movie S1 depicting the propagation of ictal multi-unit action potentials across part of a Utah array. The black arrows show multiple contiguously active areas that are separated by a mid-range mm-sized distance, supporting that the excitatory axon collateral connections are invoked for propagation of the ictal activity. Color scale represents the number of spikes per second.

Not all patients with implants in recruited territory showed spatiotemporal patterns resembling a sinc-function, and the clinical etiologies for these patients may offer some clues about why this is the case. The diffuse depressions observed in the spatial domains for both Patients 4 and 5 (Fig. 3D, S2C, S2D, E) are consistent with a local flood of excitation. Indeed, the seizures in both of these patients were characterized as secondarily generalized. This suggests that in generalized seizures, the mid-range excitatory connectivity structure (as represented by the sinc-function) may play a diminished role in comparison to other mechanisms of ictal propagation, such as local excitation or engagement of white matter tracts (Fig. 6A). Furthermore, a unique case is Patient 3, who was diagnosed with cortical dysplasia. The STA is sinc-like, and the st-SCA partially resembles a sinc-function (Fig. S2C). Cortical dysplasias have been shown to be associated with functional connectivity defects (Hong et al., 2017; Jeong et al., 2014; Rezayev et al., 2018) which may explain the partial donut ring of activity in the st-STA (Fig. S2C).

From these results, we propose that focal seizures that engage mid-range excitatory circuits may be identified by their spike-LFP spatiotemporal patterns. With this information, clinicians can potentially target specific mechanisms underlying a patient’s seizures and choose appropriate therapeutic strategies. For example, removal of horizontal interactions on a mm-scale has been the rationale for performing subpial transections in patients with intractable epilepsy (Morrel et al., 1989). In these cases, characterization of the st-SCA may inform the appropriateness of such interventions in personalized patient treatment plans.

In addition, our results suggest that these spatiotemporal patterns may be obtained without the use of MEAs. While MEAs are advantageous for monitoring and studying seizure activity with high temporal and spatial resolution, their current clinical utility is limited as they cannot be easily used to sample from multiple cortical areas. Our theoretical model, however, showed that a sinc-function in the temporal domain can predict the presence of a donut-shaped ring of activity in the spatial domain.

Consequently, a clinician could hypothesize that mid-range excitatory connections may be involved during a patient’s focal seizure if a sinc-function is observed in the temporal domain. Interestingly, we found that the sinc-function can be characterized in the temporal domain by using spiking and LFP information from a random subset of only eight electrodes (Fig. S7). This suggests that the st-SCA may be characterized by using neocortical microelectrodes that allow for recording from multiple areas by reducing the number of channels per probe. The development of such electrodes is technologically feasible as similar probes already used clinically for the monitoring of deep brain structures (Misra et al., 2014).

## Materials and Methods

### Patients

Seven patients with pharmacoresistant focal epilepsy underwent chronic intracranial EEG studies to help identify the epileptogenic zone for subsequent removal. Patients 1, 4, 6, and 7 were recruited at Columbia University Medical Center, and Patients 2, 3, and 5 were recruited from Massachusetts General Hospital/Brigham and Women’s Hospitals (Table S1). Procedures were approved by the Internal Review Board committees at Columbia University Medical Center, The University of Chicago Comer Children’s Hospital, and Massachusetts General Hospital/Brigham and Women’s Hospitals. The patients’ surgeries and treatment plans were not directed by or altered as a result of these studies.

### Signal acquisition and pre-processing

A 96-channel, 4 × 4mm MEA (Utah array; Blackrock Microsystems) was implanted along with subdural electrodes (ECoG) with the goal of recording from seizure onset sites. Additional details of study enrollment and surgical procedures have been previously published (Schevon et al., 2012; Truccolo et al., 2014). Signals from the MEA were acquired continuously at a sample rate of 30 kHz per channel (0.3-7500Hz bandpass, 16-bit precision, range ±8 mV). The reference was epidural. Up to three seizures from each patient were selected for detailed analysis to avoid biasing the dataset from the patients from whom many seizures were recorded. Seizure recordings were categorized as recruited or unrecruited territory using previously described methods (Schevon et al., 2012). Channels and time periods with excessive artifact or low signal-to-noise ratio were excluded.

Unit activity was identified using filtered 0.3-3kHz signals with spikes defined as deflections ≥4 standard deviations below the mean. The low frequency component of the local field potential (LFP) activity across the array was created by averaging the artifact-free LFP activity from all micro-electrode signals filtered 2-50Hz. The averaged LFP procedure has been shown to generate signals that are representative of and comparable to nearby electrocorticography signals (Eissa et al., 2017; Eissa et al., 2018).

### Spatiotemporal spike-centered average (st-SCA) calculations and signal analysis

All signal processing and statistical analyses were performed in MATLAB (MATLAB, Natick, MA, USA). The st-SCA was determined as follows. First, as described in (Eissa et al., 2017; Eissa et al., 2018), we detected the spikes in the multi-unit activity. Next, we collected the spatiotemporal data around each spike and translated the time and position of all associated LFPs relative to each spike’s time and position (Fig. 1). Finally, we summed all translated data and computed the average at each time and position by dividing the sum by its number of contributions. Note that this position-dependent average is necessary because not every position receives the same number of contributions during the translation of the LFP’s axes.

Evidence of radial symmetry of the st-SCA (Fig. 3C) allowed conversion from Cartesian coordinates (*ξ, ψ*) coordinates to polar coordinates (*r, θ*). By ignoring the minor deviations from radial symmetry, we focused on the spatial component of the st-SCA with respect to *r* (Fig. 5A), which enabled us to depict the spatiotemporal properties in two dimensions (Fig. 5). Furthermore, if we compute the sum across space, we obtain purely the temporal component of the st-SCA, which is equivalent to the STA. Similarly, summation over time *τ* generates the spatial component of the st-SCA. With these results, we can assess to what extent our model of the ictal network, a linear time-invariant (LTI) system with unit impulse response *C*(*τ*) *∝ sinc*(*r, τ*), fits the data. A stepwise description of this method can be found in the Supporting Information.

For calculations involving the spatial filtering of LFP signals, we applied the spatial whitening process as described in Hyvärinen et al. (2001) and Telenczuk et al. (2017). As previously published, a signal is spatially filtered by matrix multiplication with a whitening matrix ***W***, where ***W*** is the inverse square root of the signal’s covariance matrix, ***C***:

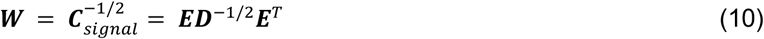

where ***E*** is a matrix of eigenvectors of ***C***_*signal*_,and ***D*** is a diagonal matrix with inverse square roots of eigenvalues *λ*_*i*_on its diagonal, such that 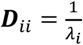 and ***D***_*ij*_ (Telenczuk et al., 2017). In this study, the signals being transformed were the MEA channel signals bandpass filtered at 2-50Hz.

## Data Availability

The data used for this study are available on reasonable request from the corresponding author (W.v.D). The data is not publicly available due to HIPAA protections, and all data sharing will have to follow protocols compliant with HIPAA policies.

## Acknowledgements

We thank Drs. Mark Kramer, Stephan A. van Gils, Hil Meijer, Douglas R. Nordli Jr., and Jack D. Cowan for valuable discussion and suggestions. C.A.S., and W.v.D. were supported by NIH Grants R01 NS095368 and R01 NS084142. C.A.S, S.S.C and E.S. were partially supported by NIH Grant R01 R01NS110669. S.L and S.S.D. were supported by University of Chicago MSTP Training Grant T32GM007281.

## Data Availability

The data used for this study are available on request from the corresponding author (W.v.D). The data is not publicly available due to HIPAA protections, and all data sharing will have to follow protocols compliant with HIPAA policies.

## Code Availability

All scripts and programs used to generate the results in this data will be made publicly available in a Github repository (https://github.com/sominlee14/stSCA_scripts).

