## Supplementary Materials for "Novel visualization of the spatiotemporal relationship between ictal spiking and LFP supports the involvement of mid-range excitatory circuits during human focal seizures"

**Supplementary Figures and Tables**

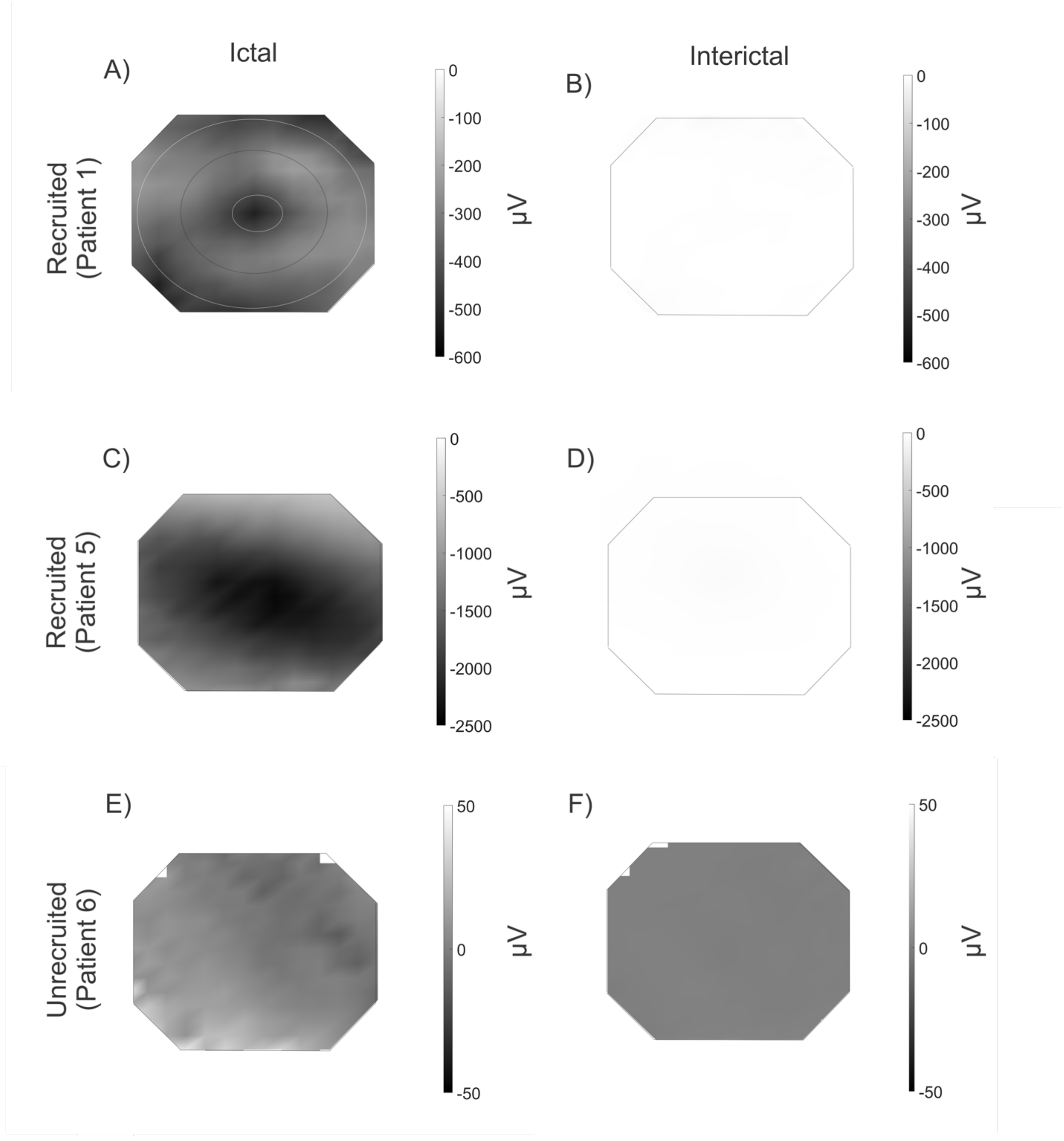


**Figure S1.** Spatial component of the spatiotemporal spike-centered averages (st-SCAs) in recruited (A—D) and unrecruited territories (E—F) show differing patterns. The st-SCA in panels A and C represent the same spike-LFP relationship as depicted in Fig. 3C and D, respectively. In all patients, the ictal signal (A, C, E) is stronger than the interictal one (B, D, F). In Patient 1 (A), the two rings surrounding the center are indicated by the circles. Patient 5 (C) instead shows a deep well of negative activity. The dynamics in unrecruited territories (E-F) are markedly different and are also much smaller in amplitude. Grayscale is in µV units

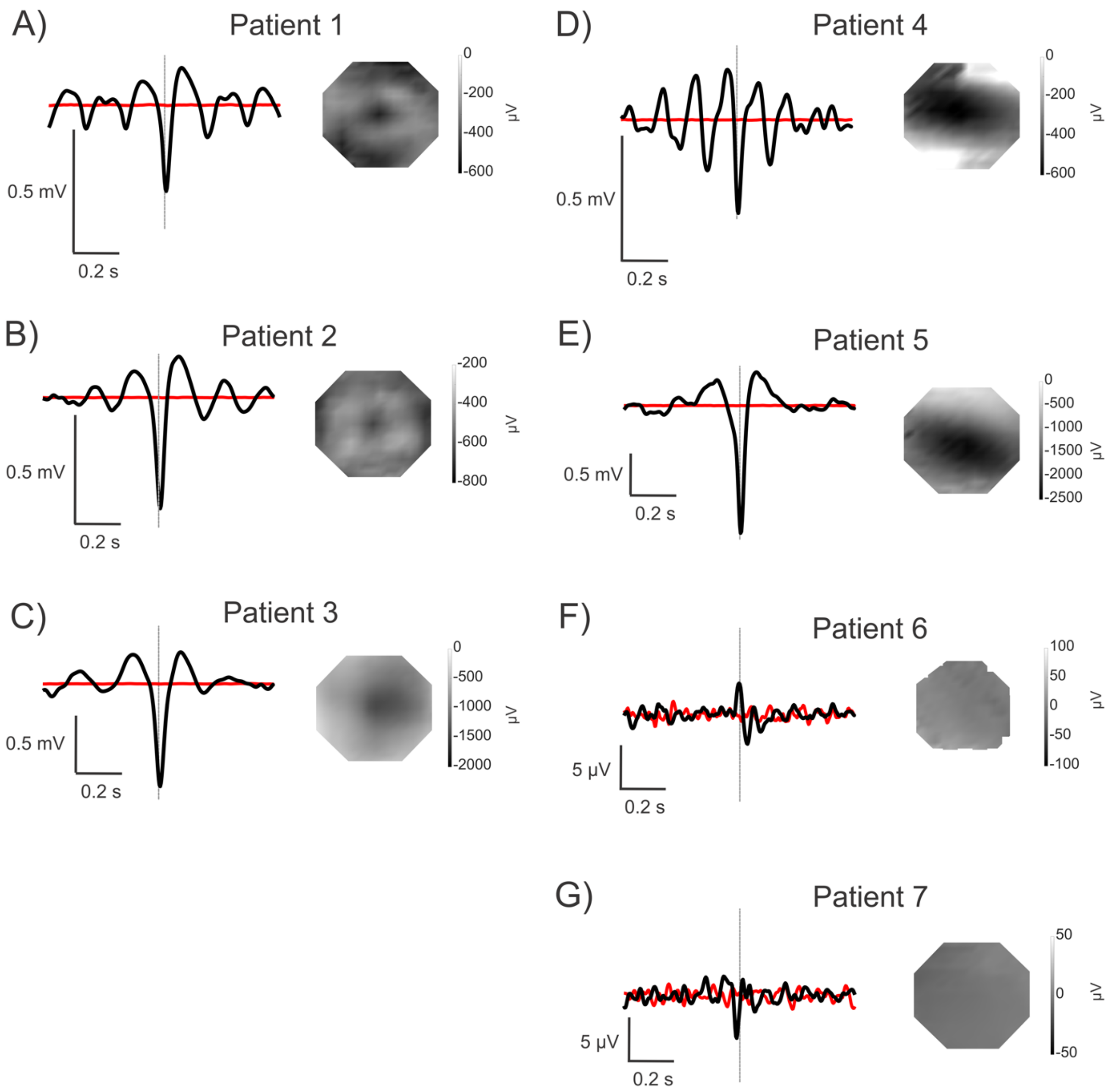


**Figure S2.** List of representative temporal and spatial components of the spike-centered averages (SCAs) for each patient. Patients 1-5 had microelectrode arrays (MEAs) implanted in recruited territory, and Patients 6-7 had MEAs implanted in unrecruited territory. Grayscale is in μV units.

A—C) Patients 1-3 resemble sinc-functions in the temporal domain.

D—E) Patients 4-5 resemble deep wells of excitatory activity in the spatial domain.

F—G) Patients 6-7 are characterized by comparatively smaller amplitude signals in both the temporal and spatial components.

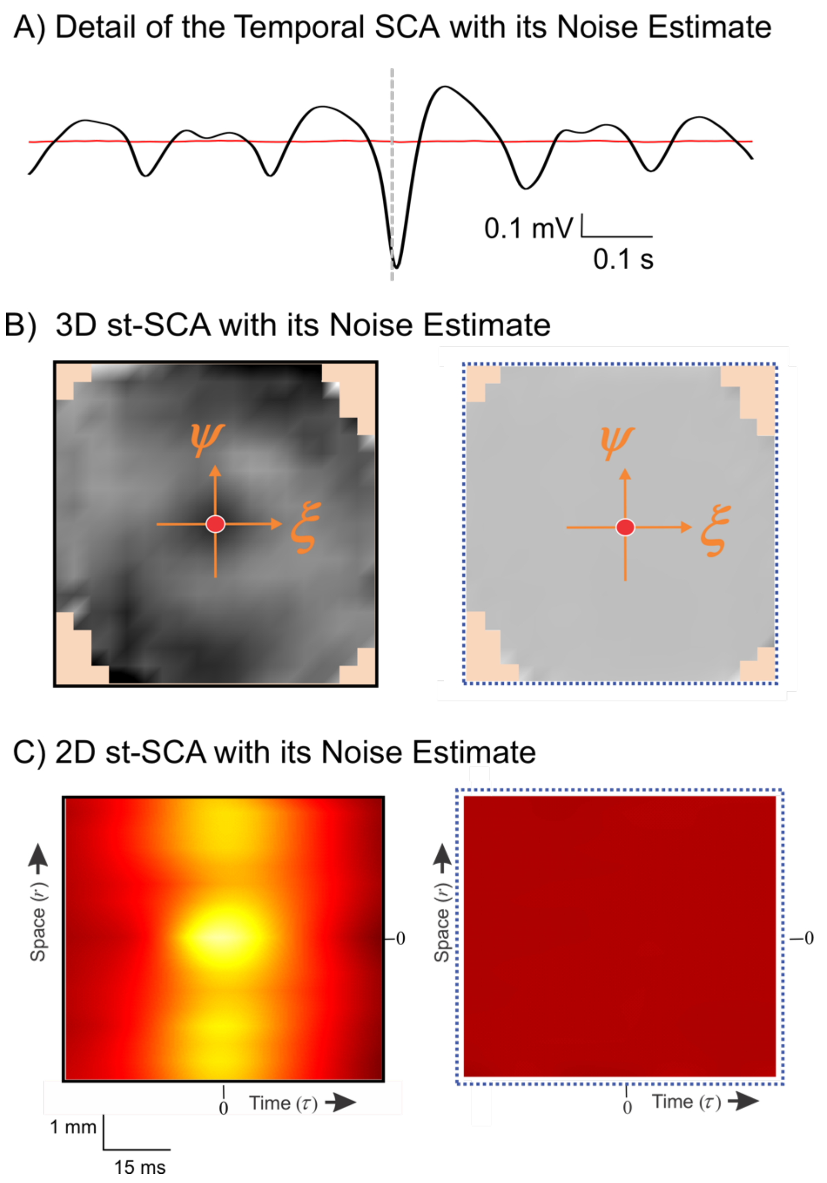


**Figure S3.** Representative noise estimates of the spatiotemporal spike-centered average (st-SCA) of Patient 1 are shown.

A) Detail of the temporal component of the st-SCA from Fig. 3A (black) and its noise estimate (red). The signal-to-noise ratio (SNR) of the depicted data is 45dB.

B) The spatial component of the st-SCA depicted in Fig. 3C (left) and its estimated noise component (right). The location specific SNR of the depicted data range is 26 – 80dB, with an average of 38dB. The units for the grayscale are identical for both maps and identical to the scale in Fig. 3.

C) The 2D st-SCA from Fig. 5C (left) and its noise estimate (right). The SNR of the depicted data is 39dB. The units for the color scale are identical for both maps and the same as in Fig. 5.

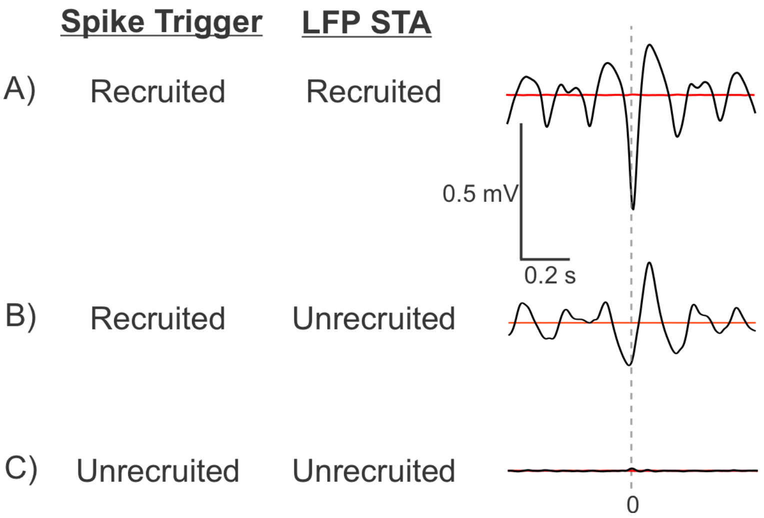


**Figure S4.** Temporal spike-triggered averages (STAs) in recruited and unrecruited territories are calculated with spike triggers from different locations. The STA of the LFP in the recruited area triggered by spikes in the recruited area (A) show a large negative peak at the time of the trigger. The STA in the unrecruited areas have a strong signal component when triggered by spikes in from the recruited areas (B), but not if triggered by spikes in the unrecruited area (C).

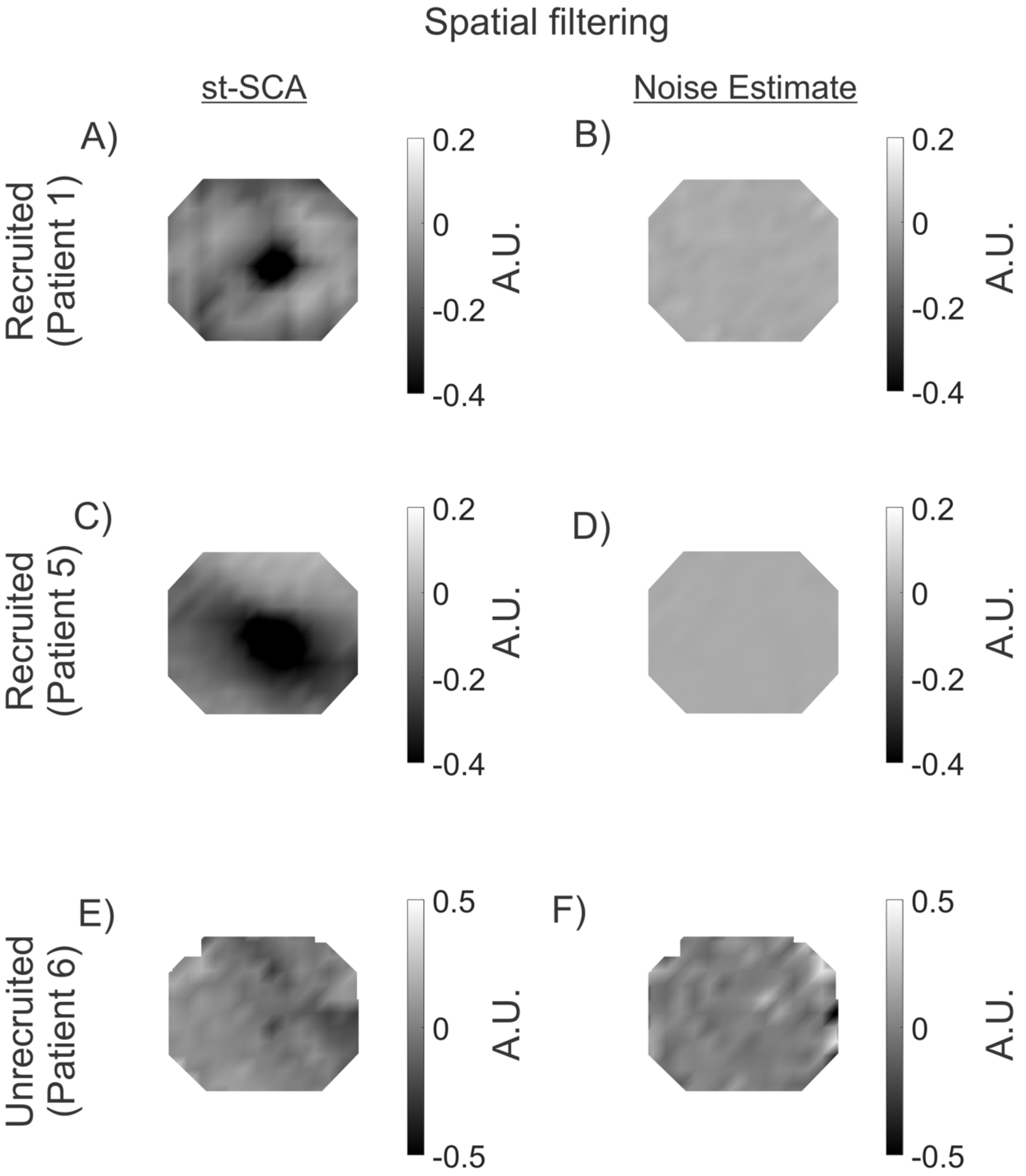


**Figure S5.** List of representative spatiotemporal spike-centered averages (st-SCAs) after spatial filtering are depicted. The spatially filtered st-SCAs resemble similar patterns to non-whitened st-SCAs, albeit a smaller amplitude signal. Grayscale is in arbitrary units (A.U.).

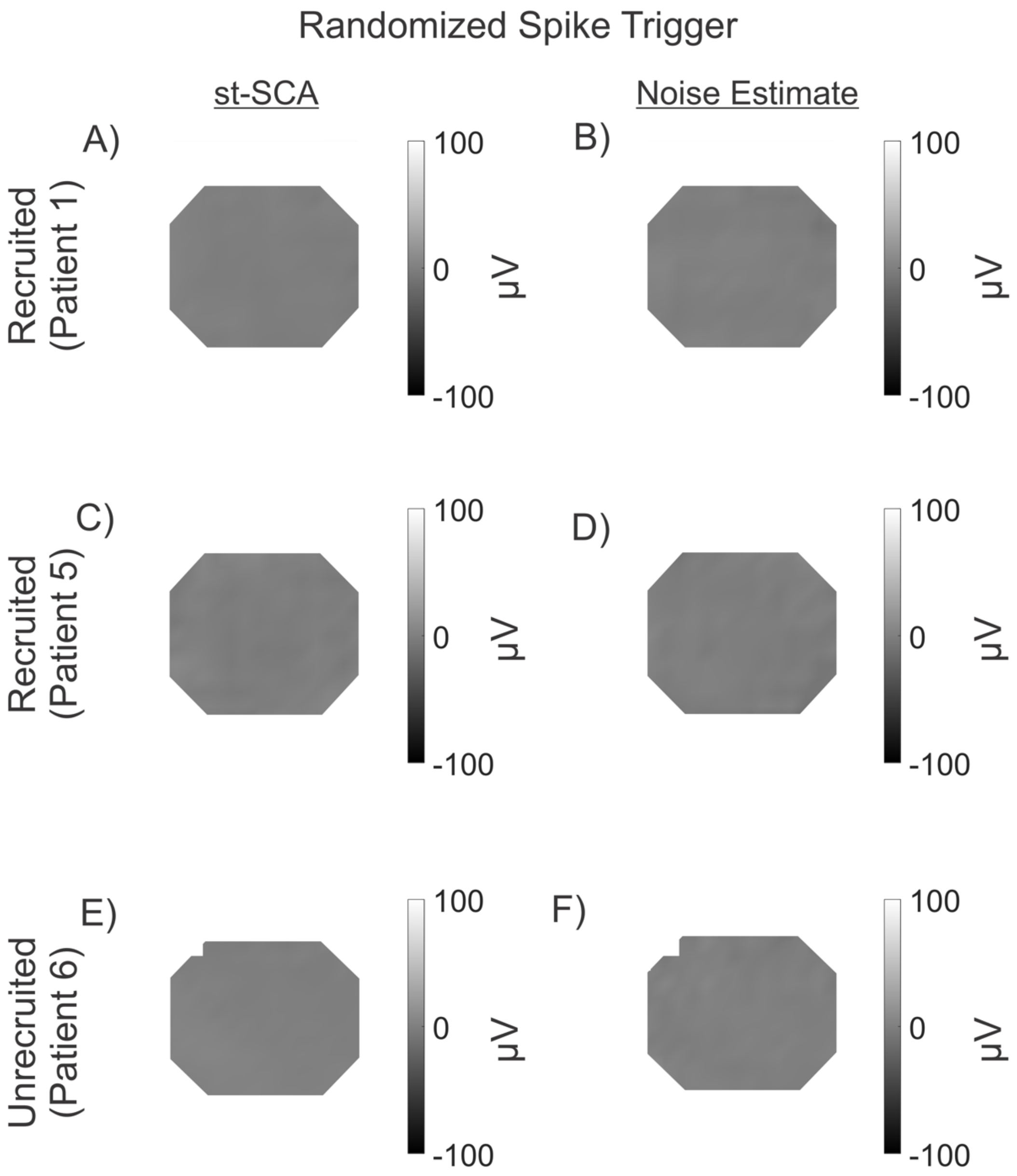


**Figure S6.** Representative spatiotemporal spike-centered averages (st-SCAs) after randomization of spike trigger timing are depicted. No spatial patterns are seen, highlighting the importance of spike timing in the st-SCA calculation. Grayscale is in μV units.

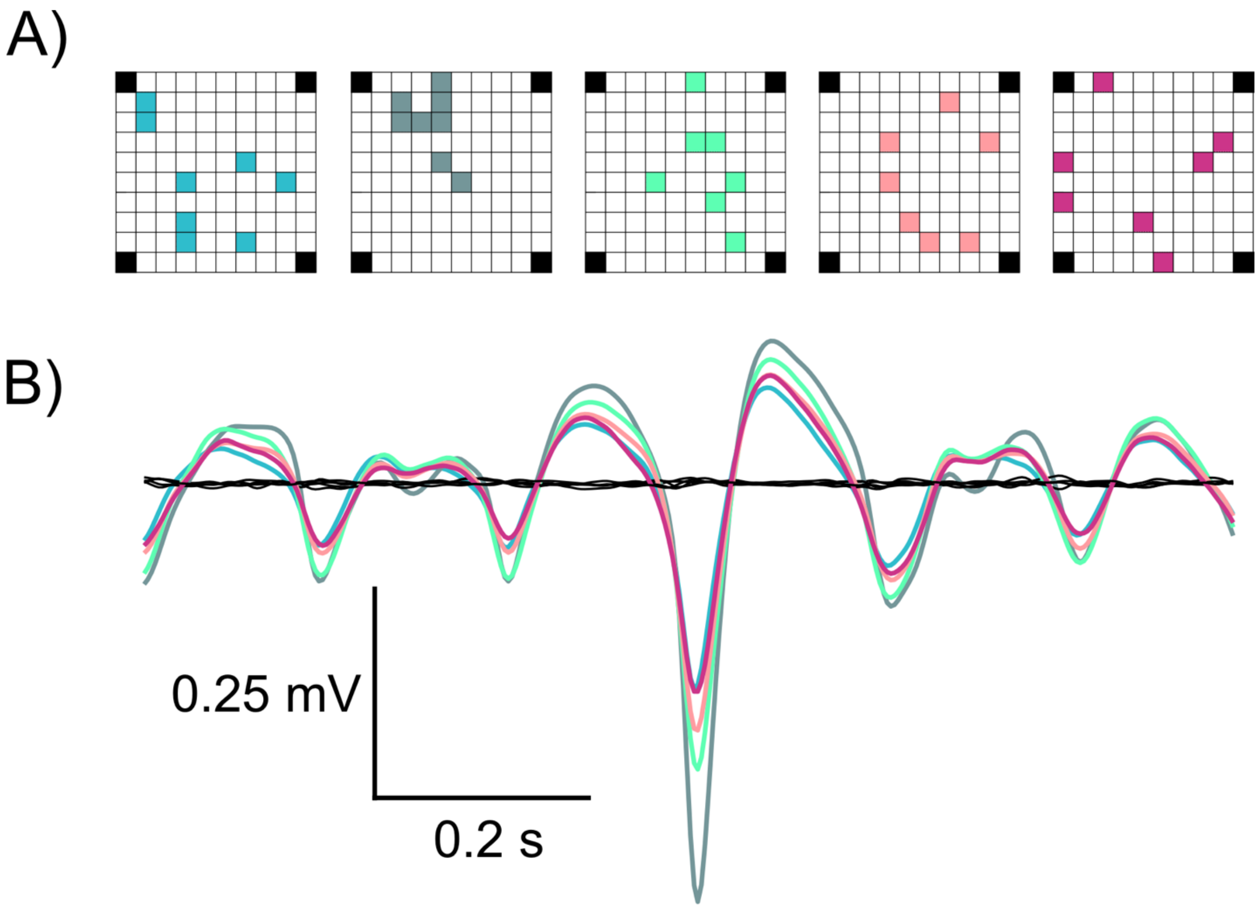


**Figure S7.** The spike-triggered average (STA) can be calculated from spike timing and LFP activity from only a random subset of eight electrodes for Patient 1. The sinc-function may be characterized in the temporal domain using signals from only eight channels across the MEA. The different colors in (A) represent different random subset of eight electrodes, and their corresponding STA in (B).

**Table S1.** Patient Table: Demographics and Clinical Features

| Patient (age/gender) | Implant location | MEA location | Seizure onset zone | No. of seizures analyzed | Seizure type(s) | Pathology |
|---|---|---|---|---|---|---|
| **Patient 1 (25yo/female)** | Left lateral and subtemporal | Left inferior temporal gyrus 2.5 cm from anterior temporal pole | Left basal/anterior temporal | 3 | Complex partial | Mild CA1 neuronal loss; lateral temporal nonspecific |
| **Patient 2 (19yo/female)** | Right lateral and subtemporal, parietal, occipital | Right posterior temporal, 1 cm inferior to angular gyrus | Right posterior lateral temporal | 1 | Complex partial with secondary generalization | Nonspecific |
| **Patient 3 (21yo/male)** | Left lateral frontal, subfrontal, temporal, subtemporal | Left middle temporal gyrus 1–2 cm posterior to the temporal tip | Left mesial temporal | 3 | Complex partial | Moderate CA3 & CA4 neuronal loss and gliosis |
| **Patient 4 (32yo/male)** | Left lateral temporal, subtemporal, parietal, frontal | Left superior temporal gyrus | Left anterior fronto-temporal | 3 | Complex partial | Cortical dysplasia |
| **Patient 5 (45yo/male)** | Right lateral temporal, parietal, frontal | Right superior temporal gyrus | Right anterior temporo-parieto-occipital | 3 | Complex partial with secondary generalization | Nonspecific |
| **Patient 6 (30yo/male)** | Left lateral frontal, mesial frontal, temporal | Left supplementary motor area, 3 cm superior to Broca’s area | Left supplementary motor area | 3 | Complex partial/tonic | N/A (multiple subpial transections performed) |
| **Patient 7 (39yo/male)** | Left lateral and mesial frontal | Left lateral frontal 2 cm superior to Broca's area | Left frontal operculum (3 × 3-cm cortical area) | 3 | Complex partial | Nonspecific |

**Table S2.** Patient Table: Seizure Recording and Spike Detection Information

| | Epoch Length (sec) | n spikes* | spikes/s* |
|---|---|---|---|
| **PATIENT 1** | | | |
| Interictal | 180 | 7720 | 43 |
| Seizure 1 | 58 | 78479 | 1353 |
| Seizure 2 | 80 | 77788 | 972 |
| Seizure 3 | 102 | 153063 | 1501 |
| **PATIENT 2** | | | |
| Interictal | 180 | 181116 | 1006 |
| Seizure 1 | 29 | 110896 | 3824 |
| **PATIENT 3** | | | |
| Interictal | 180 | 16582 | 92 |
| Seizure 1 | 52 | 162707 | 3129 |
| Seizure 2 | 88 | 274656 | 3121 |
| Seizure 3 | 57 | 193733 | 3399 |
| **PATIENT 4** | | | |
| Interictal** | 180 | 23881 | 133 |
| Seizure 1 | 82 | 385978 | 4707 |
| Seizure 2 | 102 | 366705 | 3595 |
| Seizure 3 | 96.23 | 322148 | 3348 |
| **PATIENT 5** | | | |
| Interictal | 180 | 52998 | 294 |
| Seizure 1 | 102 | 304058 | 2981 |
| Seizure 2 | 101 | 314402 | 3113 |
| Seizure 3 | 73 | 349189 | 4783 |
| **PATIENT 6** | | | |
| Interictal | 180 | 24438 | 136 |
| Seizure 1 | 12 | 3471 | 289 |
| Seizure 2 | 13 | 4902 | 377 |
| Seizure 3 | 6 | 1635 | 273 |
| **PATIENT 7** | | | |
| Seizure 1 | 20 | 17157 | 858 |
| Seizure 2 | 23 | 7778 | 338 |
| Seizure 3 | 31 | 7065 | 228 |

*Across all channels of the MEA.

**Due to limitations in available recordings, this interictal clip is 12 minutes away from the nearest known ictal activity

### Supplementary Methods

*Signal Analysis*

The spatiotemporal spike-centered average (st-SCA) was determined using the following steps (Fig. 1).

1. Each broadband signal of the 10×10 MEA was bandpass filtered for the low frequency component (2-50Hz) of the local field potential (LFP) and for spike detection (0.3–3kHz).
2. Spikes were detected in the multi-unit activity as negative deflections that exceeded four standard deviations of the filtered signal. A complete list of spike detection results can be found in Table S2.
3. For each spike the $10 \times 10$ frames of the LFP data were collected for $\pm n$ sample times representing ±5s around the spike time, and the timescale of the frames was set such that the spike occurred at time zero, $\tau = 0$.
4. All LFP frames associated with a single spike were translated such that the spike location was at the origin of the new spatial coordinate system $\xi, \psi = 0{,}0$. Note that this spatial translation is necessarily spike specific because spikes do occur at different locations.
5. Next, the translated $10 \times 10 \times (2n + 1)$ frames were put into a three dimensional $19 \times 19 \ \times (2n + 1)$ configuration with the spatiotemporal origin ($\xi, \psi, \tau = 0{,}0{,}0$) is at position $10{,}10, n + 1$. This step was done to keep the LFP frames compatible across spikes.
6. For each spike, these frames were summed into a three dimensional $19 \times 19 \times (2n + 1)$ matrix.
7. For each position in the $19 \times 19 \times (2n + 1)$ matrix, the total number of contributions $N$ was counted.
8. Finally, to obtain the spatiotemporal cross-correlation, the sum obtained in step 6 was divided by the $N$ obtained in step 7 for each position. This resulted in the discrete spatiotemporal estimate of $C(\xi, \psi, \tau)$, as shown in Eq. (3).

*Noise Estimation*

To evaluate the signal-to-noise ratio (SNR) of the averaged results, we estimated the residual noise using the plus-minus averaging approach. We implemented this by employing the above eight steps while keeping two three-dimensional 19 × 19 matrices: one summed the even contributions for each location and the other summed the odd ones. To obtain the averages for the odd and even components, each position in the matrix was then divided by its number of contributions. The sum of the even and odd averages is the same result obtained in step 8 above. In contrast, the difference between the even and odd averages cancels the consistent component (i.e., the signal) while preserving the random noise estimate (4). The $SNR$ was estimated by computing the root mean square (rms) of the signals and the rms of their noise estimates, leading to a signal-to-noise ratio, $SNR = 20 \ log\left(\frac{rms_{signal}}{rms_{noise}}\right)$ dB. Average ratios for the st-SCAs across space and time all were >30dB.

**Supplementary Text**

Computation of the spatiotemporal spike-centered average (st-SCA) using ictal recordings presents a challenge because the occurrence of action potentials across a seizing network is not experimentally controlled, unlike the scenario in which the location and timing of the neuronal activities are evoked by external stimuli. The approach as outlined in Eq. S1-S3 addresses this problem and demonstrates that the st-SCA is spatiotemporal analog of well-known spike-triggered average (STA).

For convenience, we repeat here that $(x, y, t)$ are the spatiotemporal components of the signals; $(x_i, y_i, t_i)$ are the spatiotemporal coordinates of spike $i$ and $(\xi, \psi, \tau)$ are the spatiotemporal components of the signal relative to the spike. Using a similar approach as in (Eissa et al., 2018), we now extend the model of the ictal network as a linear time invariant (LTI) system with the multi-unit action potential activity as input, the LFP as its output, and the network's unit impulse response (UIR) (see Main Text Eq. 5) defined as the LFP associated with a single unit impulse ($\delta$):

$$UIR = st\text{-}SCA \; = C(\xi, \psi, \tau) \qquad \text{(S1)}$$

We now can recover the network output $Z$ using the convolution of the $UIR$ and the network's input, i.e. the spikes:

$$Z \; = \; \iiint C(\xi, \psi, \tau) \left\{ \frac{1}{N} \sum_{i=1}^{N} \delta(x - x_i - \xi, y - y_i - \psi, t - t_i - \tau) \right\} \, d\xi \; d\psi \; d\tau \qquad \text{(S2)}$$

Note that we used the $\frac{1}{N}$ scaled version of the input here. Plugging in the expression for $C(\xi, \psi, \tau)$ results in:

$$Z = \iiint \left\{ \frac{1}{N} \sum_{i=1}^{N} LFP(x_i + \xi, y_i + \psi, t_i + \tau) \right\} \ldots$$

$$\ldots \left\{ \frac{1}{N} \sum_{i=1}^{N} \delta(x - x_i - \xi, y - y_i - \psi, t - t_i - \tau) \right\} d\xi \; d\psi \; d\tau \qquad \text{(S3)}$$

Exchange of the summation and integration operations and evaluation of the triple integral gives the model's estimate of the spatiotemporal $LFP$ from the LTI system:

$$Z = \frac{1}{N^2} \sum_{i=1}^{N} \sum_{i=1}^{N} LFP(x, y, t) \; = \; LFP(x, y, t) \qquad \text{(S4)}$$

As shown in (Eissa et al., 2018), the time domain component of this linear estimate produces a close approximation of the ongoing seizure activity with significant correlation ($p < 0.01$) between recorded and estimated activity.

*Mechanisms involved in focal seizures*

By combining current and previous findings on ictal dynamics, we can outline the following summary for an evolving neocortical focal seizure. At the micro and meso-scales, an ictal wave of action potential activity propagates at a velocity of ~1 mm/s by invoking excitation via the local connections over distances < 1 mm. This wave of hyperexcitation propagates locally when the inhibition in front of this wave fails to constrain the excitation (Eissa et al., 2017; Schevon et al., 2012; Tryba et al., 2019). In this context, it is interesting to note that this propagation process seems compatible with the evolution of the clinically observed Jacksonian march first described by Hughlings Jackson in 1870 (Extercatte et al., 2015). We now find evidence that, in addition to the slow propagation process, the ictal wave excites cortical areas farther than 1 mm away, probably via axon collaterals within the gray matter, which allows excitation to 'escape,' and enables recruitment of additional cortical territory. This activation of areas > 1 mm away might also explain modular propagation of ictal activity, a property previously observed in experimental seizures (Trevelyan et al., 2006). At the macro-scale, white matter intracortical connections are invoked, spreading ictal activity across a cm-sized territory. The activity in this macroscale territory is still highly correlated with the action potential activity in the ictal wave located in the recruited territory rather than the local action potential activity located in the non-recruited areas (Fig. S1) (Eissa et al., 2017). In addition, while local inhibition fails at the ictal wavefront, longer range inhibition remains intact and plays a critical role in sustaining the synchronous oscillatory component of the ongoing seizure at the macroscale (Eissa et al., 2017; Eissa et al., 2018).

**Legend for Supplemental Movie**

**Movie S1.** This video shows the multi-unit activity depicting the propagation of the ictal wavefront (bottom, left) and the associated 2-50 Hz low frequency component of the LFP (bottom, right) from the MEA from Patient 1. The top trace shows summed LFP activity from all MEA channels with the vertical red line noting the current time in the recording. Seizure onset is at 15 seconds, and seizure termination is at 73 seconds.
